# The role of masks in reducing the risk of new waves of COVID-19 in low transmission settings: a modeling study

**DOI:** 10.1101/2020.09.02.20186742

**Authors:** RM Stuart, Romesh G Abeysuriya, Cliff C Kerr, Dina Mistry, Daniel J Klein, Richard Gray, Margaret Hellard, Nick Scott

## Abstract

**Objectives:** To evaluate the risk of a new wave of coronavirus disease 2019 (COVID-19) in a setting with ongoing low transmission, high mobility, and an effective test-and-trace system, under different assumptions about mask uptake.

**Design:** We used a stochastic agent-based microsimulation model to create multiple simulations of possible epidemic trajectories that could eventuate over a five-week period following prolonged low levels of community transmission.

**Setting:** We calibrated the model to the epidemiological and policy environment in New South Wales, Australia, at the end of August 2020.

**Participants:** None

**Intervention:** From September 1, 2020, we ran the stochastic model with the same initial conditions(i.e., those prevailing at August 31, 2020), and analyzed the outputs of the model to determine the probability of exceeding a given number of new diagnoses and active cases within five weeks, under three assumptions about future mask usage: a baseline scenario of 30% uptake, a scenario assuming no mask usage, and a scenario assuming mandatory mask usage with near-universal uptake (95%).

**Main outcome measure:** Probability of exceeding a given number of new diagnoses and active cases within five weeks.

**Results:** The policy environment at the end of August is sufficient to slow the rate of epidemic growth, but may not stop the epidemic from growing: we estimate a 20% chance that NSW will be diagnosing at least 50 new cases per day within five weeks from the date of this analysis. Mandatory mask usage would reduce this to 6–9%.

**Conclusions:** Mandating the use of masks in community settings would significantly reduce the risk of epidemic resurgence.

## Introduction

There is increasing evidence that localized suppression or even elimination of COVID-19 is possible with interventions such as physical distancing, lockdowns, travel restrictions, testing, tracing, and quarantine. This outcome was very likely achieved in several countries, including New Zealand, Iceland, Taiwan, Thailand, and Vietnam, among others [1]. However, even if the epidemic has been locally controlled, new outbreaks can emerge if community transmission has not been eliminated and cases escape detection or quarantine, or if infected people arrive from abroad or interstate and interact with the local community (as recently seen in cities such as Melbourne and Auckland). In settings with low numbers of active COVID-19 infections, minimizing the risk of epidemic resurgence is essential for sustainability. Therefore, it is crucial to identify and quantify strategies to reduce this risk.

In this paper we focus on New South Wales, Australia’s most populous state with 7.5 million residents, as an example of a setting with low transmission, high mobility, and a well-functioning test-and-trace system. After an initial wave of COVID-19 infections in March and subsequent lockdown in April, New South Wales began relaxing physical lockdown measures over May and was experiencing near-zero case counts by the start of June, with students back at school, businesses reopening and social/community activities resuming. In late June several clusters of new infections were detected, which subsequently led to a two-month long period of low but steady case counts (between 5–20 newly detected cases per day). This experience contrasts sharply with that of the neighboring state of Victoria, which had also achieved near-zero case counts by early June but which then experienced a large second wave, with 14,434 new cases detected between 14 June and 14 August, 90% of which have been traced back to just four index cases [2].

The dynamics of COVID-19 transmission are complex, and in low-transmission settings the probability of maintaining epidemic control depends on numerous factors outside of policy control, including the characteristics of people who get infected: the size of their households, the type of work that they do, and a number of other socio-economic factors that may influence their contact networks, access to testing and capacity to self-isolate. Several studies have pointed to the role of superspreading events and overdispersion of infections in COVID-19 transmission [3–4], including work by our group examining the Seattle epidemic found that infections are overdispersed, with ∼60% of infected people not transmitting at all, while 9% cause half of all onward transmission [5], study on the role of superspreading events. As a result, even with physical distancing, high levels of testing, and rapid contact tracing, there is still a non-zero probability that a sustained outbreak could occur depending on who gets infected and where.

There are numerous non-pharmaceutical interventions (NPIs) that jurisdictions can adopt to improve their resilience to renewed epidemic waves whilst still allowing social and economic activity to continue. Such NPIs include physical distancing regulations, hygiene protocols, and the use of face masks, all of which have been recommended by the World Health Organization (WHO) [6]. These NPIs are supported by a growing body of evidence regarding their efficacy in preventing individual-level COVID-19 transmission [7–11]. However, Australia’s response through to the end of August focused on the first two measures, with only Victoria having mandated the use of face masks – and even then only after the second wave of infections was well underway and lockdown measures were in place – while other jurisdictions have only encouraged mask use in particular settings.

In this work, we assess the likelihood of epidemic rebound following a prolonged period of low, stable transmission and relatively high community mobility, using a stochastic agent-based mathematical model of COVID-19. Because the model is stochastic, we can run it multiple times and evaluate the probability of observing a given outcome among all possible outcomes. We use this feature to evaluate the probability that New South Wales will experience an epidemic resurgence under different assumptions about the adoption of masks/face coverings.

## Methods

### Demographics and networks

We began by simulating a population representative of New South Wales by taking data on the age and sex composition of the population from the 2016 census (the latest available), and using it to create a model population of agents with similar characteristics. The simulations consist of 100,000 individual agents, who are dynamically scaled based on prevalence to represent the total New South Wales population of 7.5 million. The dynamical scaling means that whenever the proportion of susceptible agents falls below a threshold of 5%, the number of agents in the model is increased; further implementation details can be found in Section 2.3.6 of Kerr et al [12].

Next, we created contact networks for these agents. The governmental response to COVID-19 in New South Wales consisted of a set of highly context-specific policies covering individuals, businesses, schools, and other types of organizations. To model these policies, we allow agents in the model to interact over five types of contact network: households, schools, workplaces, and static and dynamic community networks. The static community network consists of interactions with friends, colleagues, or other known associates who come together on a regular and predictable basis, and contains four sub-networks: professional sports, community sports/fitness/leisure clubs, places of worship, and socializing with friends. The dynamic community network consists of interactions in which people interact with strangers or random groups of people, and contains seven separate sub-networks, representing: (1) arts venues such as museums, galleries, theatres, and cinemas, (2) large events such as concerts, festivals, sports games, (3) pubs and bars, (4) cafes and restaurants, (5) public parks and other outdoor settings, (6) public transport, (7) all other community settings. The method for constructing these networks is described in our previous study of the Victorian epidemic [13] and is based on the methodology of the SynthPops Python package [14].

### Disease transmission model

We used an agent-based microsimulation model, Covasim [12], developed by the Institute for Disease Modeling and previously adapted by our group to model the Victorian epidemic [13]. Covasim contains detailed descriptions of age-dependent disease acquisition and progression probabilities, duration of disease by acuity, and the effects of interventions including symptomatic and asymptomatic testing, isolation, contact tracing, and quarantine, as well as other NPIs such as physical distancing, hygiene measures, and protective equipment such as masks. Importantly, it also captures individual variability, with viral loads varying both between individuals and over time.

### Modeling interventions and policy restrictions

Throughout March, the policy response to COVID-19 in New South Wales progressed from guidelines encouraging precautionary handwashing and distancing to a much more restrictive “lockdown” phase, in which people were only allowed to leave their houses for a limited number of reasons. This phase was maintained throughout April and then gradually eased over May–July.

Figure 1 presents a summary of how contact networks and the relative risk of COVID-19 transmission in different settings changed as policies evolved. Some of these changes in transmission risk are derived from available data [15], while others are taken from a similar modeling exercise conducted in Victoria, in which a panel of Australia-based experts reviewed the likely effect of policies on transmission risks [13]. Further details of all policies and how we model their effects on transmission risk are contained in Supplementary Table 1. Most relevantly for our subsequent analyses, we assume that the proportion of New South Wales residents who wore masks in dynamic community settings increased over August to reach 30% by the end of the month.

**Figure 1.**
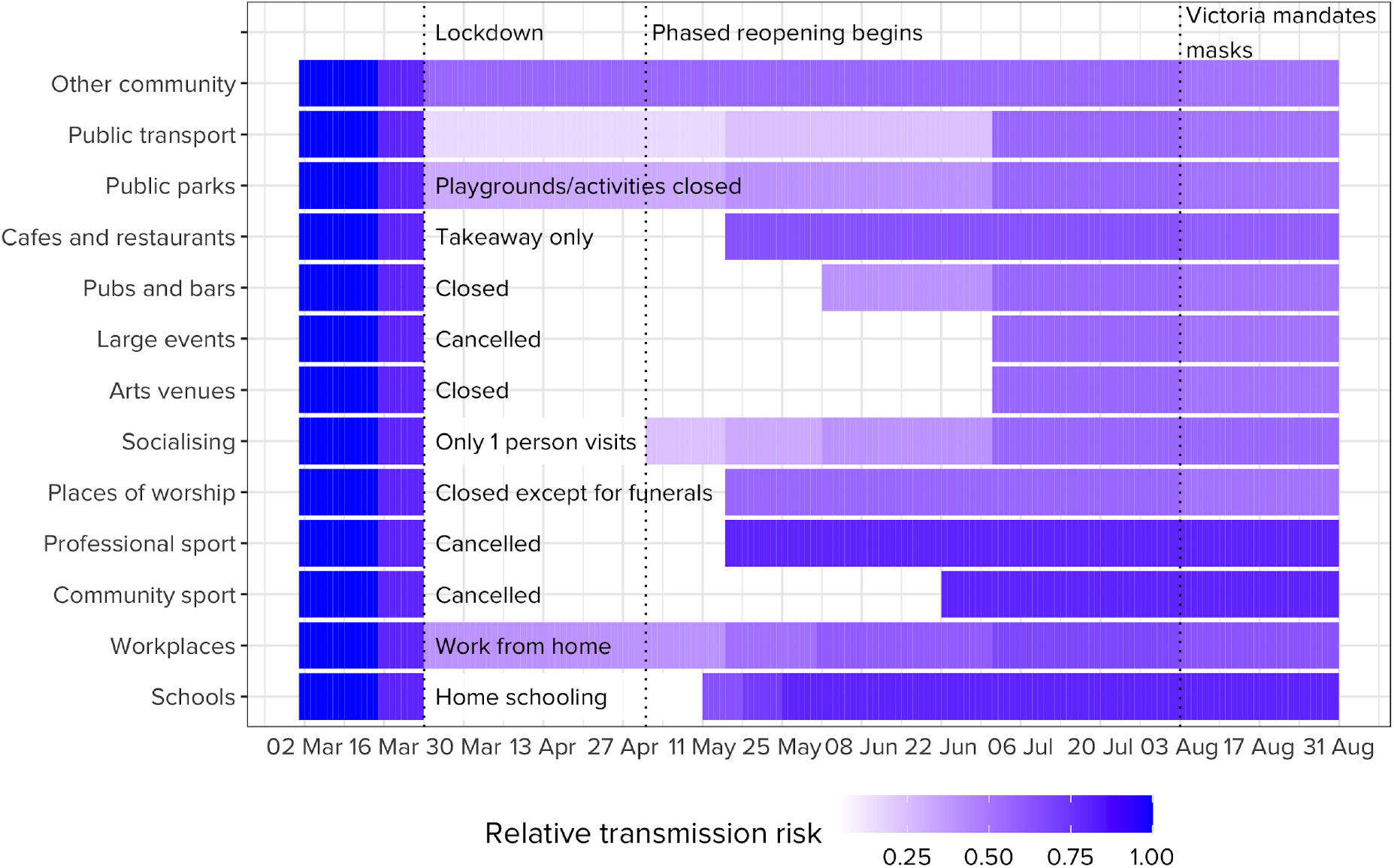
Relative changes in network structure and transmission risk across different settings in New South Wales over March–August. The absolute transmission risk varies by setting and is highest in household and lowest in outdoor settings (see [13] for details).

### Data and calibration

We initialized the model on March 1, 2020 by seeding 100 infections in the model population, with the number of seed infections chosen as part of the calibration process. The model was calibrated to data on (1) the number of tests conducted and (2) the daily number of cases diagnosed in NSW, excluding cases acquired overseas, by performing an automated search for the values of the per-contact transmission risk and the number of seed infections that minimized the absolute differences between the model projections and the data. We repeated the initialization 100 times, each time with a different set of 100 people infected at the beginning of the simulation.

Figure 2 displays the outcome of this, with the model capturing the initial outbreak, the decline in cases following the April lockdown, and then the gradual increase in June/July as policies eased, new cases arrived from interstate, and new clusters began to form. By the end of August, we estimate just over 500 active infections in New South Wales.

**Figure 2.**
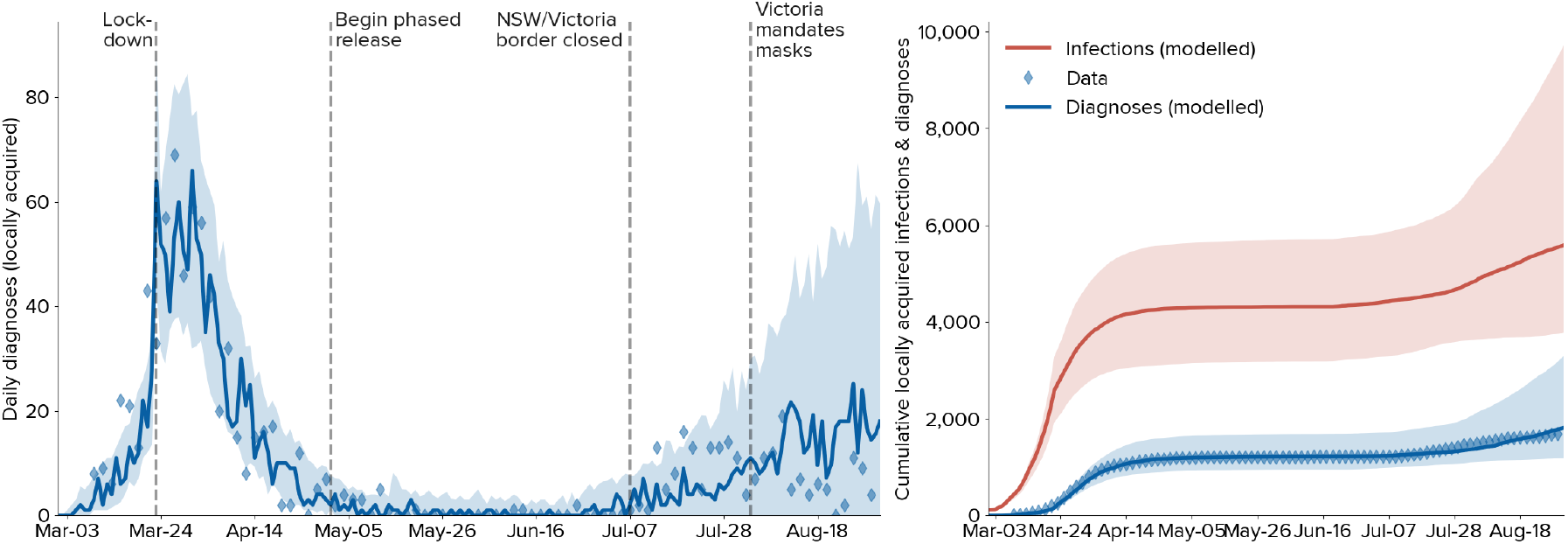
Fitting a microsimulation model to the NSW epidemic. Solid lines indicate the median model projections over 100 model runs; shaded areas indicate 95% projected intervals over different initializations; blue diamonds indicate data on confirmed locally-acquired cases.

### Model analyses

We use the model to investigate the probability of a setting with low transmission and high mobility experiencing a resurgence in cases, grounding the analysis based on the policy settings and epidemic state of New South Wales at the end of August. To calculate this probability, we reinitialize the model on August 31, 2020 and project forward by five weeks using the parameter values obtained via the calibration process, and beginning with the estimated epidemic state on August 31, 2020. We repeat this 100 times, with each iteration representing a different realization of the possible future transmission dynamics. We then calculate the proportion of simulations in which the number of cases being diagnosed per day exceeds different thresholds within five weeks.

As a baseline, we assume that the policy and behavioral settings in place at the end of August continue, including the assumption that 30% of adults wear masks while at work and when participating in community-based activities along with strangers or random groups of people. We then model two alternative scenarios:

1. No mask scenario: assuming negligible mask use;
2. Near-universal mask uptake: assuming that 95% of the adult population wear masks while at work and when participating in community-based activities along with strangers or random groups of people.

The individual-level effectiveness of masks at reducing COVID-19 transmission is difficult to determine, and will be influenced by the level of restrictions or NPIs already in place. A comprehensive meta-analysis covering 41 studies of mask effectiveness concluded that masks are associated with a reduction in infection for mask-wearers by at least one-third compared to control groups [16]. We assume that masks will reduce the per-contact probability of transmission by an additional 20% relative to a baseline in which other NPIs are in place. We also conduct a sensitivity analysis in which the individual-level effectiveness of masks is assumed to be 30%.

## Results

Beginning from a point with ongoing low levels of community transmission, high mobility, and with 30% of adults wearing masks at work and in settings with unknown groups of people, we estimate that there is a 20% chance that transmission will increase to at least 50 new cases per day within five weeks (Figure 3). If masks were not worn, the probability would increase to 25%, indicating that the current level of mask usage does not have a great impact on containing the probability of a resurgent epidemic. However, we find that under the near-universal mask uptake scenario, the probability of diagnosing more than 50 cases per day within five weeks would fall to 8% (Figure 3A). Furthermore, we estimate that the total number of infections over the next five weeks would be 20% lower than under a scenario in which masks are not used, largely driven by a 37% reduction in infections transmitted in dynamic community settings (Figure 3C). Under all scenarios, we estimate that the median number of daily infections is likely to continue to gradually increase (Figure 3D).

**Figure 3.**
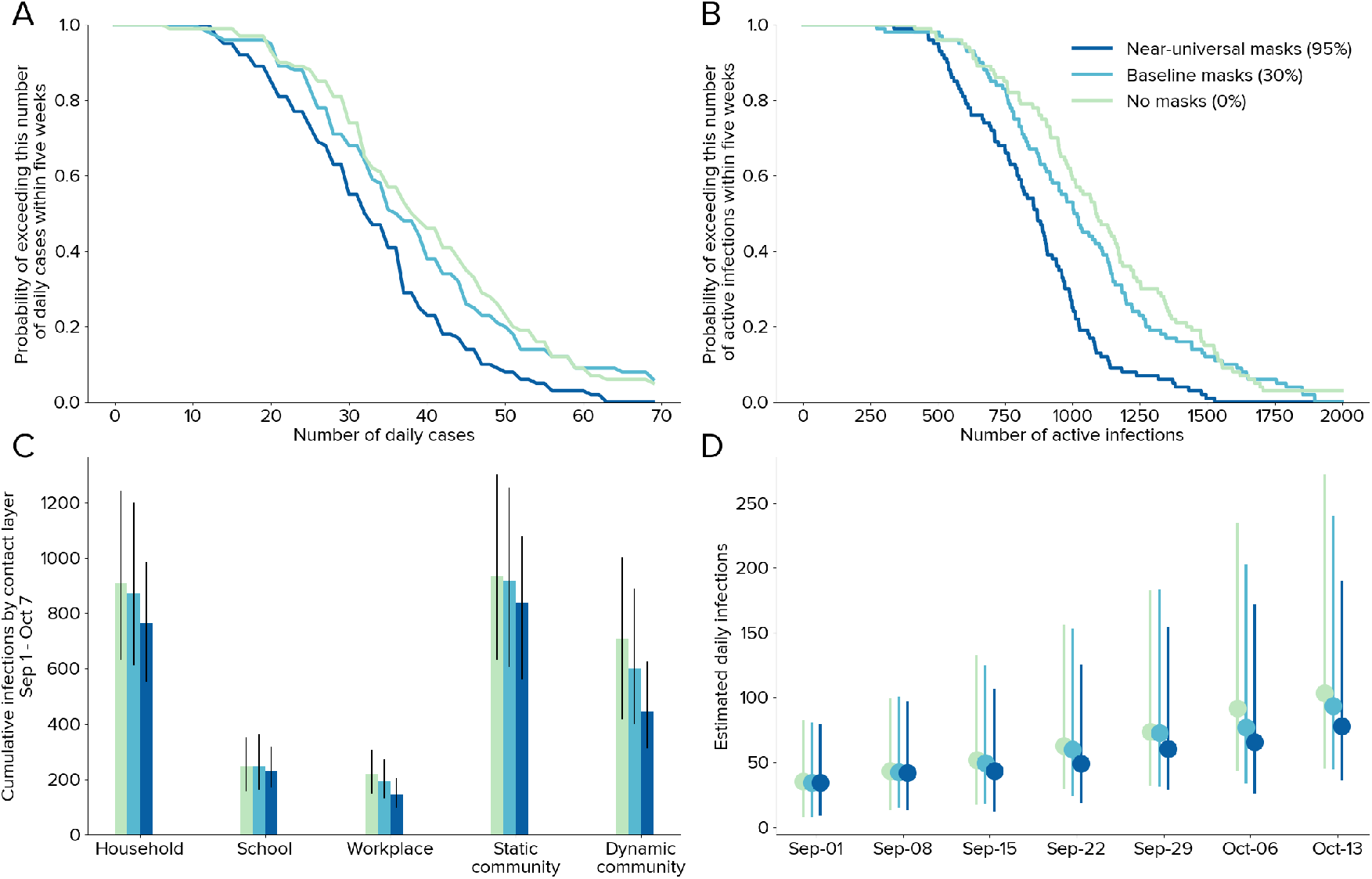
Quantifying the likelihood of epidemic resurgence in New South Wales. (A) Probability of exceeding a given number of daily new diagnoses within five weeks; (B) the probability of exceeding a given number of active infections within five weeks; (C) cumulative infections in different contexts (colored bars indicate median, black error bars indicate 95% projected intervals), showing that the use of masks leads to a first-order reduction on transmissions in dynamic community contact networks; (D) daily estimates of the number of new infections at the end of each week under the three mask scenarios (dots indicate median, colored bars indicate 95% projected interval).

We also conducted a sensitivity analysis in which individual-level mask effectiveness was assumed to be 30%. Under this scenario, we estimate an 18% probability of diagnosing more than 50 new cases per day within five weeks if mask uptake does not change, or 6% under the high mask scenario. Overall infections over the next five weeks would be 30% lower if masks were adopted with high uptake (Figure S1).

## Discussion

Evidence from numerous other settings has shown that as long as the population remains susceptible to COVID-19 infection, reopening society is likely to lead to new epidemic waves unless a well-operating test-and-trace strategy is in place [5], [17]–[19]. In this work, we examined a low-transmission, high mobility setting with limited mask usage, and found that high levels of testing and contact tracing have thus far succeeded in controlling the epidemic, but do not eliminate the risk of an epidemic resurgence. At the time of writing, case numbers in New South Wales had decreased from ∼15/day over July to < 5/day by the end of August, which may indicate that test-and-trace efforts in this instance were sufficient to curtail the risk of epidemic resurgence. However, our main finding does not depend on the exact date of the analysis; as long as viral transmission remains in the community, the probability of epidemic resurgence (e.g., an untraceable cluster, a failure of quarantine, or a superspreading event) remains.

We found that the use of masks would reduce the probability of epidemic resurgence in New South Wales: if the use of masks was mandated in New South Wales, we estimate that the probability of diagnosing at least 50 new cases per day within five weeks would reduce from 20% to 9%. Given that masks are an intervention with low social and economic costs, and containing an epidemic resurgence is extremely disruptive and costly (with Treasury estimates suggesting that Victoria’s Stage 4 restrictions will result in an AU$7–9 billion reduction to national GDP in the September quarter [20]), this reduction in risk is of major significance and suggests that a policy of mandatory mask use is likely to have an extremely high benefit-to-cost ratio.

This study adds to a sizable body of evidence supporting the adoption of face masks as a low-cost means of protecting individuals from acquiring COVID-19 [7], [8], [10], [21]. Modeling studies have shown that the population-level effects of masks depend on the state of the epidemic. A study from Israel showed that masks are particularly effective when the effective R is close to 1, and can determine whether a low-level epidemic tips into an outbreak or not [22]. In higher transmission settings where the effective R is greater than 1, two studies found that masks are most effective when used in conjunction with a collection of other NPIs [9], [23]. To our knowledge, no studies to date have examined the extent to which face masks can prevent a resurgent outbreak in low-transmission settings. Furthermore, by using a model that already incorporated the numerous other COVID-19 control measures in place in New South Wales, we illustrate that face masks have benefit even in the context of a well-functioning test-and-trace system.

There are several limitations to this study. Firstly, the mathematical model that we use is subject to the usual limitations of mathematical models, including uncertainty around the parameters that characterize COVID-19 transmission and disease progression, uncertainty around the impact of interventions and behavioral changes, and reliance on data sources (such as the number of COVID-19 cases by likely source) that may be incomplete and/or subject to revision. To the extent possible, we managed these issues by sampling parameters from probability distributions and conducting sensitivity analyses around the efficacy of masks. Secondly, we made assumptions about the proportion of contacts of diagnosed cases that can be traced within a certain number of days; further data on these proportions would greatly improve model estimates. Thirdly, our analyses assume that the policy environment in New South Wales would be relatively slow to react to an increase in case numbers; we focused on the question of quantifying the likelihood of diagnosing more than 50 cases/day on the assumption that this would equate to a high likelihood that New South Wales would enter a more restrictive phase of lockdown, but a faster policy reaction, as recently seen in Auckland, would change the nature of the results seen here.

Our work suggests that adoption of face masks by the general public could substantially reduce the risk of new epidemic waves. Given that individuals are already requested to isolate if they have been diagnosed with COVID-19 or are displaying symptoms, a major benefit of masks is in controlling asymptomatic transmission, which is estimated to make up approximately one third of all transmissions. The use of masks also has a role in reinforcing the importance of other NPIs. Not only does this have positive health outcomes in terms of reducing the number of COVID-19 infections and associated mortality, but it also has clear social and economic benefits by mitigating the need for more extreme lockdown measures that are required to curtail epidemic resurgences once they have begun.

## Data Availability

Only publicly available data were used

## Acknowledgements

The authors would like to thank additional members of the Institute for Disease Modeling team who contributed to the base Covasim model. The authors gratefully acknowledge the support provided to the Burnet Institute by the Victorian Government Operational Infrastructure Support Program. This work benefited from a useful discussion with James Wood, Deborah Cromer, and John Kaldor of the University of NSW, Australia. MH is the recipient of an Australian National Health and Medical Research Council fellowships.

## Supplementary materials

**Table S1:**
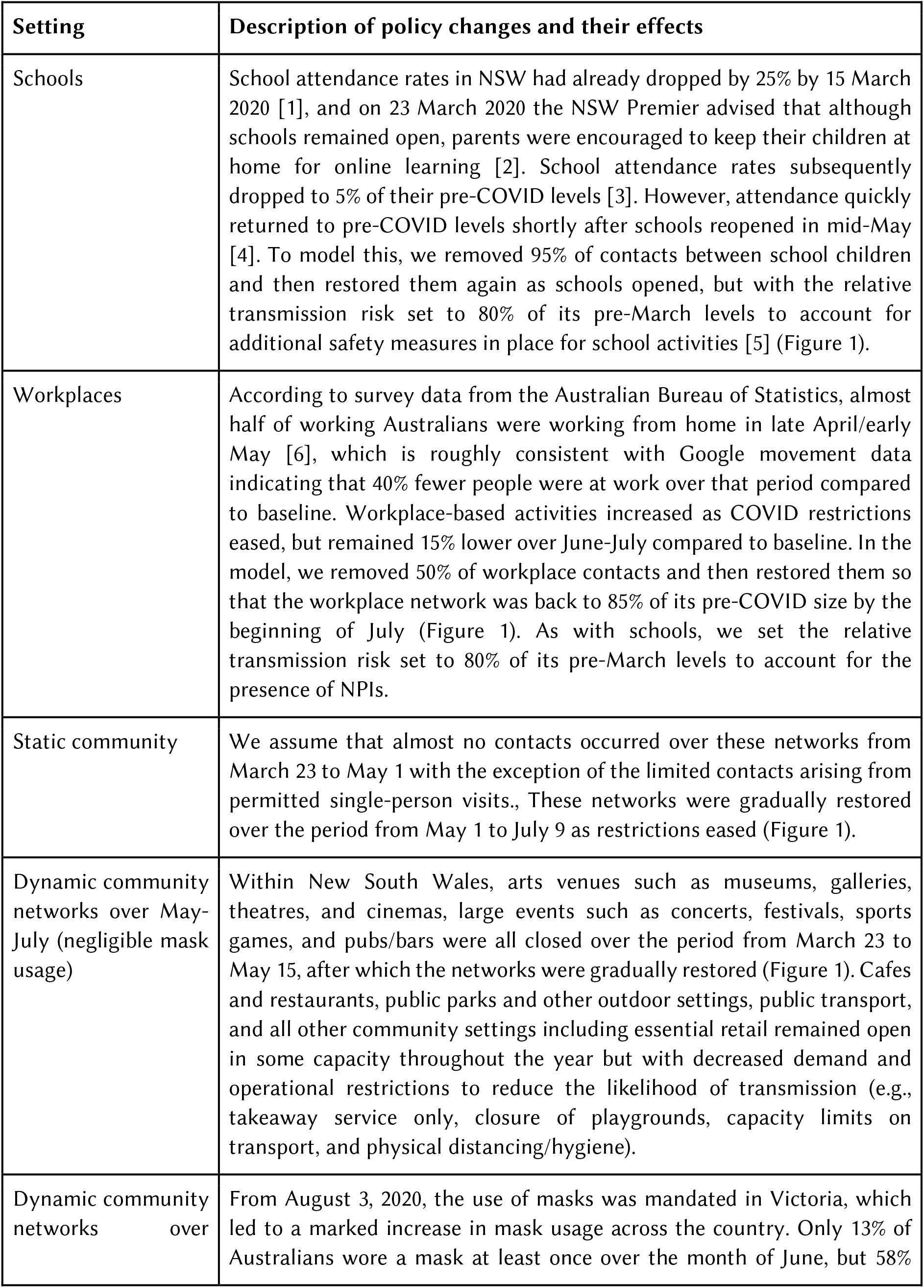
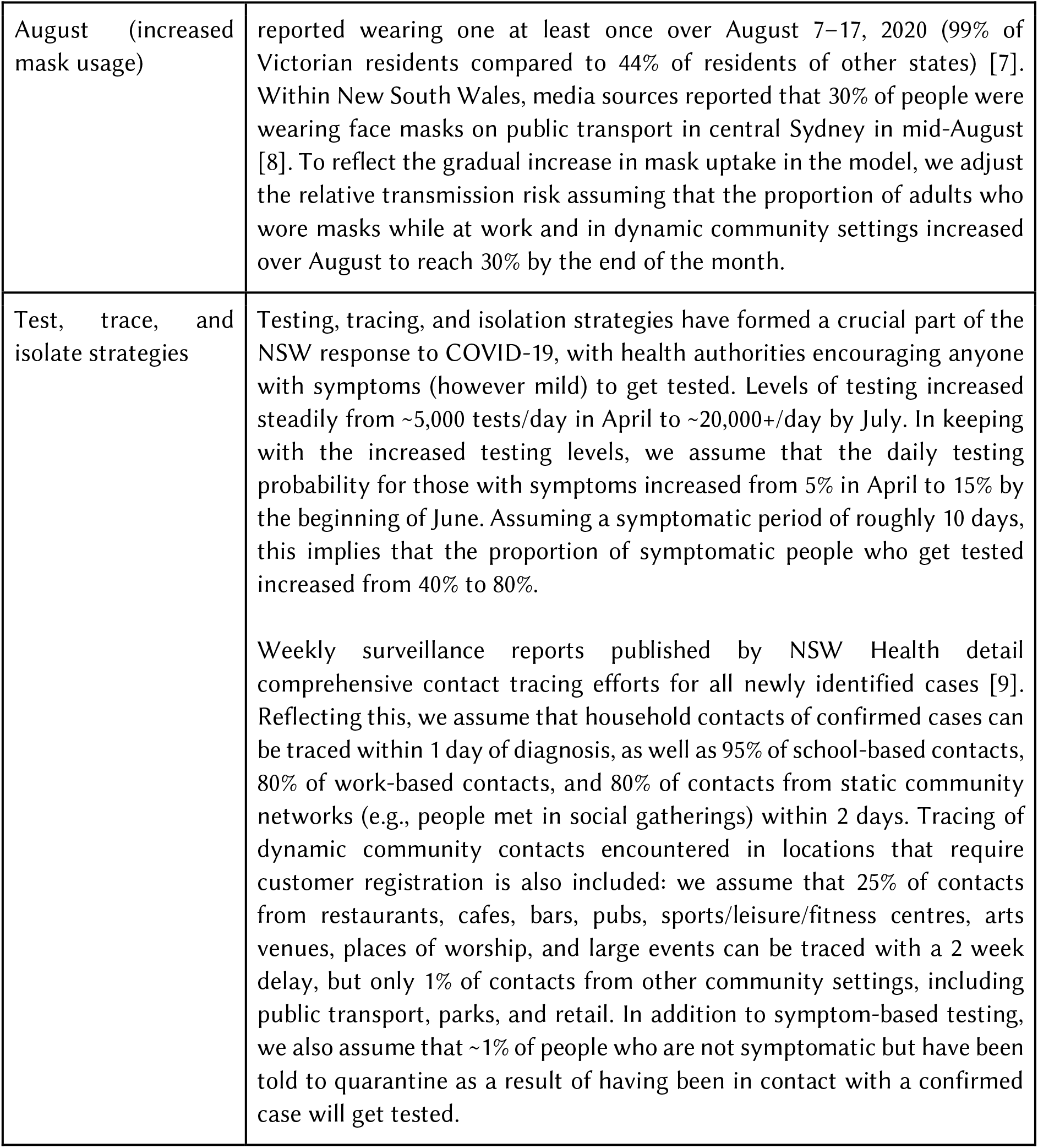
Effects of policies on transmission risk in New South Wales

**Figure S1.**
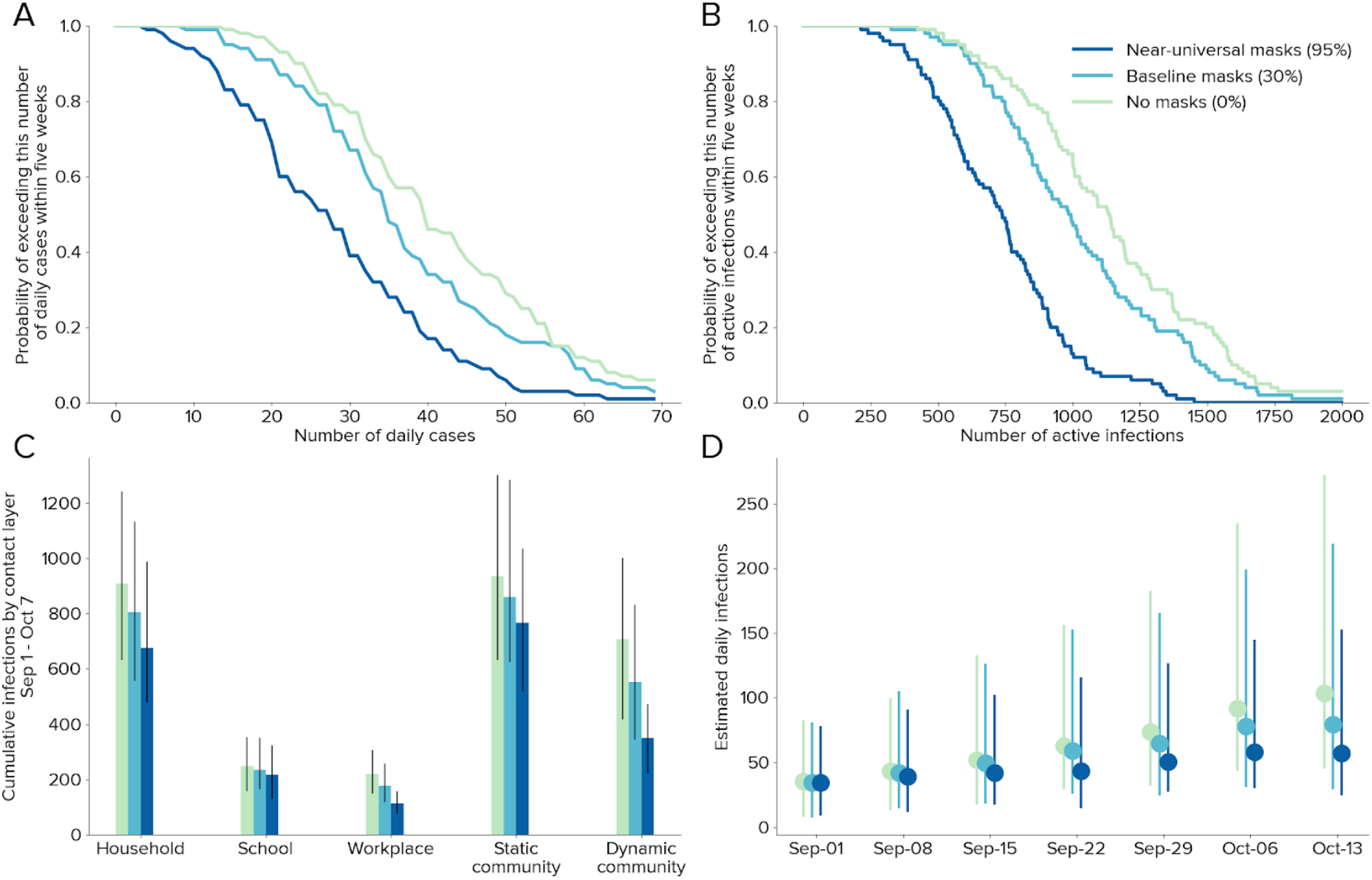
The likelihood of epidemic resurgence in New South Wales, assuming individual-level mask effectiveness of 30%

## References

[1] “Countries beating Covid-19,” EndCoronavirus.org. https://www.endcoronavirus.org/countries (accessed Aug. 19, 2020).

[2] N. Mills and T. Towell, “Family of four staying at Rydges seeded 90% of second-wave COVID cases,” The Age, 2020.

[3] A. Endo, Centre for the Mathematical Modelling of Infectious Diseases COVID-19 Working Group, S. Abbott, A.J Kucharski, and S. Funk, “Estimating the overdispersion in COVID-19 transmission using outbreak sizes outside China,” Wellcome Open Res, 5, 67, Jul. 2020, doi: 10.12688/wellcomeopenres.15842.3

[4] B.M. Althouse et al., “Stochasticity and heterogeneity in the transmission dynamics of SARSCoV-2,” arXiv:2005. 13689 [physics, q-bio], May 2020, Accessed: Aug. 29,2020 [Online]. Available: http://arxiv.org/abs/2005.13689.

[5] C.C Kerr et al., “Controlling COVID-19 via test-trace-quarantine,” 2020. https://www.medrxiv.org/content/10.1101/2020.07.15.20154765v3 (accessed Aug. 19, 2020).

[6] World Health Organization, “ Advice on the use of masks in the context of COVID-19,”; 2020. [Online]. Available: https://www.who.int/emergencies/diseases/novel-coronavirus-2019/advice-for-public/when-and-how-to-use-masks.

[7] D.K. Chu et al. “ Physical distancing, face masks, and eye protection to prevent person-to-person transmission of SARS-CoV-2 and COVID-19: a systematic review and meta-analysis,” The Lancet, 395, no.10242, pp. 1973–1987, Jun. 2020, doi: 10.1016/S0140-6736(20)31142-9.

[8] N.H. Leung et al. “ Respiratory virus shedding in exhaled breath and efficacy of face masks,” Nature Medicine, vol. 26, no. 5, Art. no. 5, May 2020, doi:10.1038/s41591-020-0843-2.

[9] S.E. Eikenberry et al., “To mask or not to mask: Modeling the potential for face mask use by the general public to curtail the COVID-19 pandemic,” Infectious Disease Modelling, vol.5, pp. 293–308, Jan. 2020, doi:10.1016/j.idm.2020.04.001.

[10] E.P Fischer, M.C Fischer, D. Grass, I. Henrion, W.S Warren, and E. Westman, “Low-cost measurement of facemask efficacy for filtering expelled droplets during speech,” Science Advances, p. >eabd3083, Aug. 2020, doi: 10.1126/sciadv.abd3083.

[11] S. Flaxman et al., “Estimating the effects of non-pharmaceutical interventions on COVID-19 in Europe,” Nature, vol. 584, no. 7820, Art. no. 7820, Aug. 2020, doi: 10.1038/s41586-020-2405-7.

[12] C.C Kerr et al., “Covasim: an agent-based model of COVID-19 dynamics and interventions,” 2020, doi: 10.1101/2020.05.10.20097469.

[13] N. Scott et al., “Modelling the impact of reducing control measures on the COVID-19 pandemic in a low transmission setting,” 2020, Accessed: Aug. 19, 2020. [Online]. Available: https://www.medrxiv.org/content/10.1101/2020.06.11.20127027v1.

[14] synthpops.org. Institute for Disease Modeling, 2020.

[15] ABC News, “Recovering from coronavirus: Data shows how Australia is rebuilding as COVID-19 restrictions ease,” 2020.

[16] IHME, “COVID-19: What’s New for June 25, 2020.”

[17] A.J Kucharski et al., “ Effectiveness of isolationisolation, testing, contact tracing and physical distancing on reducing transmission ofchapter-title>Effectiveness SARS-CoV-2 in different settings,” Epidemiology, preprint, Apr. 2020. doi: 10.1101/2020.04.23.20077024.

[18] J. Hellewell et al., “ Feasibility of controlling COVID-19 outbreaks by isolation of cases and contacts,” The Lancet Global Health, vol. 8, no. 4, pp. e488–e496, Apr. 2020, doi: 10.1016/S2214-109X(20)30074-7.

[19] M.E Kretzschmar, G. Rozhnova, M.C.J Bootsma, M. van Boven, J.H.H.M vande Wijgert, and M.J.M Bonten, “Impact of delays on effectiveness of contact tracing strategies for COVID-19: a modelling study,” The Lancet Public Health, vol. 5, no. 8, pp. e452–e459, Aug. 2020, doi: 10.1016/S2468-2667(20)30157-2.

[20] “PM suggests $9 billion cost of stage 4 restrictions in Melbourne,” Aug. 06, 2020. https://www.abc.net.au/news/2020-08-06/victoria-coronavirus-crisis-blows-out-gdp-estimates-by-billions/12530130 (accessed Aug. 26, 2020).

[21] M. Liang et al., “Efficacy of face mask in preventing respiratory virus transmission: A systematic review and meta-analysis,” Travel Med Infect Dis, p. 101751, May 2020, doi: 10.1016/j.tmaid.2020.101751.

[22] B. Javid and N.Q. Balaban, “ Impact of Population Mask Wearing on Covid-19 Post Lockdown,” Infectious Microbes & Diseases, Jun. 2020, doi: 10.1097/IM9.0000000000000029.

[23] L. Worden et al., Estimation of effects of contact tracing and mask adoption on COVID-19 transmission in San Francisco: a modeling study. https://www.medrxiv.org/content/10.1101/2020.06.09.20125831v1 (accessed Aug. 20, 2020).

## Supplementary materials references

[1] J. Baker, “Coronavirus: NSW school attendance falls 25 per cent as soap, toilet paper shortage hit” The Sydney Morning Herald, 2020.

[2] National Centre for Immunisation Research and Surveillance, “COVID-19 in schools – the experience in NSW,” 2020.

[3] E. Elsworthy, “Students are being sent back to school in NSW soon — here’s what you need to know,” ABC News, Apr. 21, 2020.

[4] N. Chrysanthos, “‘There were bubbles and balloons’: High attendance on first day back at school, ” The Sydney Morning Herald, May 25, 2020.

[5] “A guide to NSW school students for Term 3.” https://education.nsw.gov.au/covid-19/advice-for-families.html (accessed Aug. 29, 2020).

[6] Australian Bureau of Statistics, “Media Release – Loneliness most common stressor during COVID-19 (Media Release)”, May 18, 2020. https://www.abs.gov.au/ausstats/abs%40.nsf/mediareleasesbyCatalogue/DB259787916733E4CA25855B0003B21C?OpenDocument (accessed Aug. 20, 2020).

[7] c = AU; o = Commonwealth of A.ou =Australian B. of Statistics“ Media Release – COVID-19 anxiety in Victoria felt Australia-wide (Media Release)”, Aug. 31, 2020. https://www.abs.gov.au/AUSSTATS/abs@.nsf/Latestproducts/4940.0Media%20Release1August%202020?opendocument&tabname=Summary&prodno=4940.0&issue=August%202020&num=&view=(accessed Sep. 02,2020).

[8] T.R.Clun Kate Aubusson, Rachel, “CCTV monitors how many are wearing masks on trains” The Sydney Morning Herald, Aug. 13, 2020. https://www.smh.com.au/national/nsw/cctv-monitors-how-many-are-wearing-masks-on-trains-20200813-p55lh4.html (accessed Sep. 02,2020).

[9] “COVID-19 weekly surveillance reports - COVID-19 (Coronavirus)” https://www.health.nsw.gov.au/Infectious/covid-19/Pages/weekly-reports.aspx (accessed Aug. 28,2020).

